# PREVALENCE OF DEPRESSION AND ANXIETY IN COLOMBIA: WHAT HAPPENED DURING COVID-19 PANDEMIC?

**DOI:** 10.1101/2023.02.23.23286343

**Authors:** Sandra Martínez-Cabezas, Mónica Pinilla-Roncancio, Gabriel Carrasquilla, Germán Casas, Catalina González-Uribe

**Author notes:** Corresponding author: Catalina González-Uribe, E- mail. These authors contributed equally to this work (Sandra Martínez-Cabezas, Mónica Pinilla-Roncancio).

## Abstract

The COVID-19 pandemic has impacted the well-being of millions of people around the globe. During the COVID-19 pandemic, the mental health of the population was affected, which means that governments would need to implement different actions to mitigate and treat mental health disorders result of the pandemic.

This study aims to estimate the prevalence of anxiety and depression for female and male adolescents and adults in Colombia before the COVID-19 pandemic. It also aimed to estimate the potential increase of the prevalence in each group as a result of the COVID-19 pandemic in 2020. We used the Individual Registry of Health Services Delivery data from 2015 - 2021 to estimate the observed prevalence of anxiety and depression. Using the National Mental Health Survey 2015, we simulated the expected prevalence of anxiety and depression for adolescents (12 to 17 years) and adults (18 or older) from 2016 to 2020. We used an arithmetic static Monte Carlo simulation process to estimate the expected prevalence. The results of the analysis using revealed an important increase in the observed prevalence of these disorders for adults and adolescents and men and women between 2015 and February 2020. When we simulated different scenarios using the National Mental Health Survey and estimated the prevalence of both depression and anxiety for adults and adolescents, we found that the prevalence of depression and anxiety has had an important increase in the last five years for all groups and had an important increase during the 2020. This increase has been greater for women than for men, and for adolescents than adults. Our results show the number of people who need potential attention from the health system in Colombia and highlight the importance to think about how to avoid and detect potential cases of anxiety and depression especially in female adolescents.

## Introduction

The COVID-19 pandemic has impacted the well-being of millions of people around the globe (1). Throughout 2020-2021, different countries worldwide implemented strict quarantine measures to prevent the spread of the virus and avoid the saturation of health services. Lockdown measures, reduction of social interactions, restrictions to carry out physical activity, exposure to traumatic situations, such as loss of family members and friends, and constant exposure to information (e.g., recommendations, prohibitions, and misinformation) were elements associated with increased risk of developing mental health problems for individuals with and without a history of mental health problems (2–4).

Most countries around the globe restricted the provision of health services to cover the extra demand of patients with COVID-19 symptoms. This change in the supply of healthcare services reduced the access that the general population had to services related to non-communicable diseases including mental health services, modifying the access to diagnosis, treatment, and the continuity of care health services, especially in low-income and middle-income countries (LMICs) (5–7).

A survey conducted by the World Health Organization (WHO) in 130 countries showed that between June and August 2020 mental health services for vulnerable populations were affected. For example, disturbances were observed in 67% of psychological counselling services and psychotherapy; 30% of the countries reported difficulties in accessing medications for the treatment of mental disorders; and mental critical health services were disrupted in 93% of countries (8).

According to the WHO, approximately one billion people in the world live with a mental health disorder, yet the provision of mental health services is one of the most neglected and underinvested areas in LMICs (9). During the COVID-19 pandemic, the mental health of the population was affected, which means that different actions will be required to prevent mental health disorders and provide access to mental health services, especially in LMICs. Different studies have estimated the potential increase in the prevalence of anxiety and depression as a result of the pandemic; however, most of them have based their figures on non-representative data. Therefore, it is not possible to know the increase in the prevalence of these disorders as a result of the COVID-19 pandemic(10,11). Aiming to provide better estimators, Santomauro and collaborators (12) analyzed the change in the prevalence of major depressive disorders and anxiety disorders before and during the COVID-19 pandemic using data from surveys in a systematic literature review. According to the results of this study, it is estimated an increase of 53.2 million additional cases of major depression, and an additional 76.2 million cases of anxiety disorders worldwide. Using this information, the WHO estimated that anxiety and depression disorders might have increased by 25% during the COVID-19 pandemic, with women and children as the most affected groups (13).

The COVID-19 pandemic notably affected countries of the Americas, it was one of the regions with the highest number of cases and deaths in the world (14), and Colombia was one of the most affected countries in Latin America and the Caribbean. Indeed, more than five million cases and more than 120,000 deaths were reported by the end of December 2021(15) and, the pandemic has generated negative impacts on the delivery of health services and the mental health of the population. The pandemic led to the formulation of strict quarantines and health services restrictions favouring elective consultation by telehealth, limiting health promotion and disease prevention activities (16,17).

Colombia estimated the prevalence of any mental disorders before the COVID-19-pandemic using the National Mental Health Survey in 2015. This survey is the only nationally representative that has estimated the prevalence of mental disorders in Colombia. The prevalence for adolescents aged 12-17 years, 12 months prior to the survey, was 4.4% (95% CI 3,3-5,7), for adults aged 18 years or older was 4.0% (95% CI 3,5-4,6). In addition, mental health disorders were more prevalent in rural areas and among people living in poverty. (18).

To analyse the potential effect of the COVID-19 pandemic on the mental health of individuals in Colombia, some studies were conducted in 2020 and 2021 by different authors. One of the first analyses estimated the prevalence of mental health disorders using a web survey between April and May 2020 (19). According to this study, 18% of the participants reported living with a mental health disorder, or that someone in their family was diagnosed with a mental health disorder in the previous six months to the survey; women and individuals aged 18 to 29 were more affected by mental disorders in comparison with men and people from other age groups. Other researchers have also identified that women, young and poor people are more vulnerable to mental health disorders (20,21). In addition, PSY-COVID-19, a study conducted in 30 countries using Patient Health Questionnaire-2 and Generalized Anxiety Disorder-2 instruments, found that 35% of respondents have a higher risk of depression, and 29% have a higher risk of anxiety (22); this percentage is lower compared to the one found by Colombia’s National Planning Department, where 52% of households reported a deterioration in mental health in adult members (23).

Studying mental health in Colombia is important, given that it is a country that has suffered an armed conflict for more than 50 years, and this situation has a direct impact on the emotions, thoughts, and mental health of its population (24) and the COVID-19 pandemic further altered mental health indicators in the country. As it was mentioned above, it is estimated that the prevalence of individuals with mental health disorders such as depression or anxiety increased in Colombia as a result of the COVID-19 pandemic; however, beyond descriptive studies, based on non-probabilistic samples, there are no studies to our knowledge that analyzed this topic There has not been a study updating the prevalence of mental health disorders in the country and the last figures are based on the National Mental Health Survey from 2015. Therefore, this study aims to: 1) provide a national estimation of how much the prevalence of mental health disorders has changed from 2015 to 2021 according to national health registers (observed prevalence), and 2) estimate different scenarios on how the prevalence of depression and anxiety might have changed during the COVID-19 pandemic in 2020 (expected prevalence).

## Materials and Methods

### Data sources

#### National mental health survey

We used data from the National Mental Health Survey 2015 a survey representative at the national and regional levels of the country. The survey excluded people aged 65 years or older with a positive dementia screen using the Mini-Mental State Examination, individuals over 12 years of age with cognitive limitations, institutionalized individuals, and those who did not speak Spanish (18). The final sample included in this study was 1754 adolescents aged 12 to 17 years and 10870 adults 18 years and older. For the effects of this study, we excluded children from 7 to11 years old because the information was provided by the primary caregiver.

#### Individual Registry of Health Services Delivery

The Individual Registry of Health Services Delivery (IRHSD) is the administrative register of healthcare providers in Colombia. It includes information on providers, consultations, and the provision of different services. Its main objective is to support the billing process of healthcare providers; therefore, the information is partial and probably does not include all providers or services. However, it is the only national register that includes information on the real use of healthcare services in the country. Thus, even though it is a limited data source, it is commonly used to analyse the demand for healthcare in the country (25).

The IRHSD includes the provision of services, classified according to ICD-10 diagnosis (International classification of diseases 10th). Information on the date of consultation, age, sex, region of residence, and type of service for everyone that has contact with the system (26), therefore, it allows the analysis of the number of outpatient consultations by ICD-10 diagnosis, according to the type of service received.

We used IRHSD data from 2015 to 2021 to estimate the observed prevalence of anxiety and depression, with the assumption that everyone who consulted was included in the register. We analyzed out-patient and in-patient (hospitalization and emergencies) cases disaggregated by type of diagnosis, sex, and age. The download date of the IRHSD was June 14^th^, 2022 and we used the number of people who attended because it is equivalent to the number of cases, given that the system computes singular people.

### Statistical analysis

#### Observed Prevalence

To estimate prevalence from 2015 to 2021, we used the total number of cases reported in the IRHSD with ICD-10 codes F41.1-F41.2-F41.3-F41.8-F41.9 for anxiety and F32.0- F321- F322- F323- F328- F329- F330- F331- F332- F333- F334- F338- F339- F341 for depression. As denominator, we used the population projections by the National Administrative Department of Statistics (DANE. From the Spanish “Departamento Administrativo Nacional de Estadística” by sex and groups of age for 2015 to 2021 (27), and we computed 95% confidence intervals. To estimate the percentage change in prevalences used this formula: ((*V*_*2*_*-V*_*1*_*)/V*_*1*_) *100, where *V*_*1*_ represents initial value and *V*_*2*_ represents the final value.

##### Simulation (Expected Prevalence)

Using the National Mental Health Survey 2015, we simulated the expected prevalence of anxiety and depression for adolescents (12 to 17 years) and adults (18 or older) from 2016 to 2020. We used two primary sources of information to inform the decision modelling. First, the estimated prevalence of anxiety and depression in Colombia reported by Santomauro (12), and second, the observed prevalence of depression and anxiety that we calculated using the IRHSD for each year between 2016 and 2020. We used an arithmetic static Monte Carlo simulation to estimate the expected prevalence. Other researchers have used these simulations in the context of mental health service planning (28,29).

We first calculated the prevalence of depression and anxiety for adults and adolescents using the National Mental Health Survey (2015). We used questions that aimed to identify individuals who had suffered any depression or anxiety disorder during the 12 months before the data was collected. Using this information as a baseline (before the COVID-19 pandemic), we calculated the rate of relative annual growth (the difference between year 2 minus year 1 divided by year 1) of the prevalence of anxiety and depression using the IRHSD for adolescents, adults, men, and women, and we computed the prevalence of depression and anxiety for the years 2016 to 2019, assuming that the observed relative annual growth in IRHSD as the real relative annual growth of the prevalence of depression and anxiety.

To analyse the potential effect of the COVID-19 pandemic on the prevalence of depression and anxiety, we estimated three different scenarios for 2020 (Figure 1):

**Figure 1.**
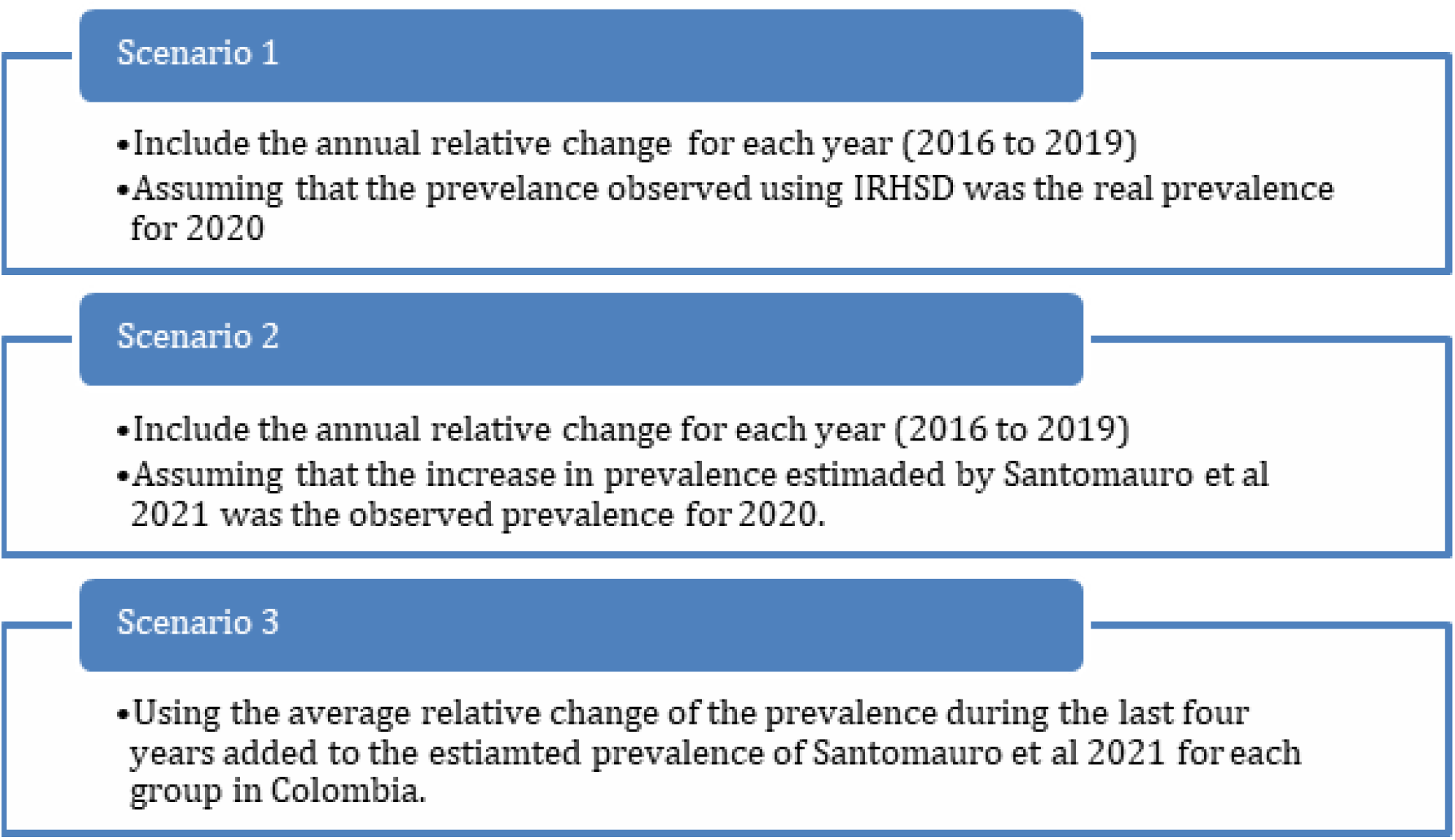
Scenarios used for the simulation of the prevalence of depression and anxiety

1. We assumed that the relative annual growth observed in IRHSD for 2020 was the real relative annual growth for 2020 (Scenario 1),
2. We used the average growth for depression and anxiety estimated by Santomauro (12); during the COVID-19 pandemic for each group (men and women, adolescents, and adults) in Colombia (Scenario 2).
3. We assumed that the prevalence of depression and anxiety would have continued to change at a similar rate to the previous years. Therefore, we used the average percentage change in the prevalence between 2016 and 2019 to estimate the annual growth for 2020 and added to this percentage, the estimated increase in the prevalence presented by Santomauro (12) for Colombia (Scenario 3).

We computed each of the three scenarios for four groups: men 12 to 17, women 12 to 17 and men 18 years or older and women 18 years or older. For all the models we computed a simulation of 1000 iterations and calculated standard errors and confidence intervals. We used Stata 17® for all the analyses.

## Results

### IRHSD: Cases

Cases of both anxiety and depression in the IRHSD show similar trends between 2015 and 2018. In 2019, the number of anxiety cases increased, and this continued during January and February 2020 that accounted for more than 61,000 cases in each month, before the pandemic was declared. However, in March 2020 when the government established the national quarantine, the number of cases went down to 47,000 and increased again at the end of the first national lockdown in September 2020, to 66,000. In January 2021 the number of cases decreased to 31,000. Similar trends were observed in the case of depression, with a decrease from 48,000 cases in January 2020 to 31,000 cases in March 2020. In September 2020 (after the national lockdown), the number of cases increased to 37,000, and decreased again to 16,000 in January 2021, situation that might be associated with the implementation of local lockdowns (See Figure 2).

**Figure 2.**
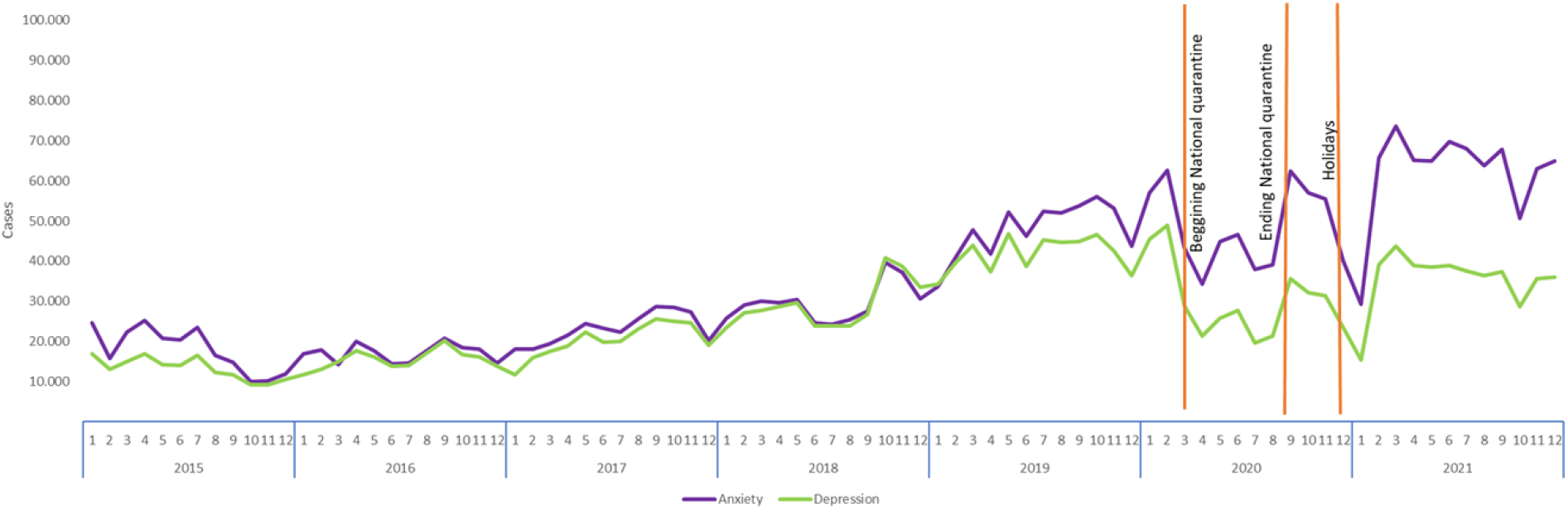
Number of cases per anxiety and depression disorders 2015-2021 by year and month in Colombia, all ages, all sexes. Source: Author’s own elaboration using data from the Individual Registry of Health Services Delivery 2016-2021

#### Observed prevalence (using the Individual Registry of Health Services Delivery)

##### Anxiety

We observed an increase in prevalence since 2015, larger in female adolescents (0.12% in 2015 to 1.46% in 2021) and adult women (0.98% in 2015 to 2.61% in 2021). In all groups, except in adult males, during 2020, there was a reduction in the prevalence of anxiety. It is important to highlight that during 2021, the increase in the prevalence was higher than 50% in adolescents and more than 10% in adults, compared to 2020. See Tables 1 and 2 for details.

**Table 1.** Prevalence of anxiety and depression- Adolescents 12-17 years old

| Year | Women |  |  |  | Men |  |  |  |
| --- | --- | --- | --- | --- | --- | --- | --- | --- |
|  | Anxiety | Annual % change | Depression | Annual % change | Anxiety | Annual % change | Depression | Annual % change |
| 2015 | 0.12 |  | 0.05 |  | 0.14 |  | 0.07 |  |
| 2016 | 0.12 | 2.24 | 0.08 | 57.34 | 0.14 | -0.77 | 0.12 | 67.49 |
| 2017 | 0.20 | 71.98 | 0.16 | 109.29 | 0.20 | 44.40 | 0.14 | 21.30 |
| 2018 | 0.33 | 61.70 | 0.36 | 118.67 | 0.28 | 36.43 | 0.23 | 65.76 |
| 2019 | 0.70 | 112.79 | 0.84 | 135.13 | 0.49 | 79.24 | 0.46 | 94.64 |
| 2020 | 0.65 | -6.86 | 0.67 | -20.37 | 0.39 | -21.26 | 0.31 | -30.93 |
| 2021 | 1.46 | 123.84 | 1.37 | 106.49 | 0.62 | 60.50 | 0.49 | 56.06 |

Source: Author's own elaboration using data from the Individual Registry of Health Services Delivery

**Table 2.** Prevalence of anxiety and depression - Adults 18 + years old

|  | Women |  |  |  | Men |  |  |  |
| --- | --- | --- | --- | --- | --- | --- | --- | --- |
| Year | Anxiety | Annual % change | Depression | Annual % change | Anxiety | Annual % change | Depression | Annual % change |
| 2015 | 0.98 |  | 0.75 |  | 0.42 |  | 0.29 |  |
| 2016 | 0.91 | -6.36 | 0.84 | 12.11 | 0.39 | -7.92 | 0.32 | 8.90 |
| 2017 | 1.16 | 27.16 | 1.06 | 25.94 | 0.52 | 33.21 | 0.41 | 30.21 |
| 2018 | 1.40 | 20.25 | 1.42 | 34.11 | 0.68 | 31.99 | 0.62 | 49.17 |
| 2019 | 2.18 | 55.90 | 1.95 | 36.78 | 1.04 | 52.28 | 0.83 | 34.12 |
| 2020 | 2.14 | -1.65 | 1.35 | -30.46 | 1.07 | 2.98 | 0.59 | -28.09 |
| 2021 | 2.61 | 21.53 | 1.46 | 7.75 | 1.21 | 13.50 | 0.63 | 6.19 |

Source: Author's own elaboration using data from the Individual Registry of Health Services Delivery

##### Depression

The prevalence of depression also had an increase between 2015 and 2021. In both, men and women and in depression and in anxiety there is an increase up to 2019, then decrease in 2020 and an important increase in 2021. Female adolescents had the largest increase from 0.05% to 1.37%. A similar pattern occurred for adult women with an increase in depression of 1%. However, in this group (adult women) the largest prevalence was in 2021, with almost 3% of women aged 18 or older reporting cases of depression. For men (adolescents and adults), the prevalence of depression increased, but not in the same proportion as for women. Adolescent men there was an increase of 0.42 percentage points between 2015 and 2021 In adults the increase in 2021 was not as evident as for adolescents. (See Tables 1 and 2).

#### Emergency and hospitalizations cases

We explored the number of cases in emergencies and hospitalizations to analyse the trends in the use of these services. As expected, we found similar trends to the ones for outpatient cases during 2020 in emergency cases between the beginning of the COVID-19 pandemic in March and the ending of the national quarantine on August 31^st^, 2020. The number of hospitalization cases decreased in March 2020 and rose in October 2020 for both, anxiety and depression (See Appendix 1 for more details).

### Estimated Prevalence: Monte Carlo Simulation

According to the Mental Health Survey, in 2015, the prevalence of anxiety was 2.3% and 1.6%, for adult women and men, respectively. In the case of female and male adolescents, the prevalence of anxiety was higher in women (4.9%) than men (2.1%) (Table 3). In addition, the prevalence of depression in adults was higher for women (2.2%) compared to men (1.0%). It is important to highlight that in the case of male adolescents the prevalence of depression was 0.7% in 2015 (Table 4).

**Table 3.** Estimated prevalence of anxiety in each simulated scenario.

| Group | Base Line 2015 | Increase 2020 by Lancet | Percentage change according to IRHSD 2020 | Estimated result for 2020 | SD | 95% Confidence Interval |  | Estimated Population | Population 2020 |
| --- | --- | --- | --- | --- | --- | --- | --- | --- | --- |
| Scenario 1 |  |  |  |  |  |  |  |  |  |
| Women (18 years or older) | 2.3% | 33.2% | -1.7% | 4.9% | 0.2% | 4.917% | 4.925% | 924,253 | 18,781,581 |
| Men (18 years or older) | 1.6% | 28.0% | 3.0% | 3.4% | 0.2% | 3.417% | 3.425% | 591,702 | 17,296,667 |
| Female adolescents (12 to 17 years) | 4.9% | 76.0% | -6.9% | 12.6% | 1.0% | 12.549% | 12.588% | 298,868 | 2,377,905 |
| Male adolescents (12 to 17 years) | 2.1% | 59.0% | -21.3% | 4.02% | 0.5% | 4.011% | 4.031% | 99,405 | 2,472,245 |
| Scenario 2 |  |  |  |  |  |  |  |  |  |
| Women (18 years or older) | 2.3% | 33.2% | -1.7% | 6.8% | 0.3% | 6.847% | 6.858% | 1,287,053 | 18,781,581 |
| Men (18 years or older) | 1.6% | 28.0% | 3.0% | 5.3% | 0.3% | 5.260% | 5.272% | 910,822 | 17,296,667 |
| Female adolescents (12 to 17 years) | 4.9% | 76.0% | -6.9% | 23.8% | 1.7% | 23.767% | 23.830% | 565,908 | 2,377,905 |
| Male adolescents (12 to 17 years) | 2.1% | 59.0% | -21.3% | 6.9% | 0.8% | 6.882% | 6.912% | 170,506 | 2,472,245 |
| Scenario 3 |  |  |  |  |  |  |  |  |  |
| Women (18 years or older) | 2.3% | 33.2% | -1.7% | 7.8% | 0.3% | 7.806% | 7.818% | 1,467,180 | 18,781,581 |
| Men (18 years or older) | 1.6% | 28.0% | 3.0% | 6.2% | 0.4% | 6.176% | 6.189% | 1,069,342 | 17,296,667 |
| Female adolescents (12 to 17 years) | 4.9% | 76.0% | -6.9% | 32.3% | 1.8% | 32.317% | 32.384% | 769,274 | 2,377,905 |
| Male adolescents (12 to 17 years) | 2.1% | 59.0% | -21.3% | 9.0% | 0.9% | 8.953% | 8.987% | 221,752 | 2,472,245 |

Source: Author's own elaboration using data from the National Mental Health Survey 2015

**Table 4.**
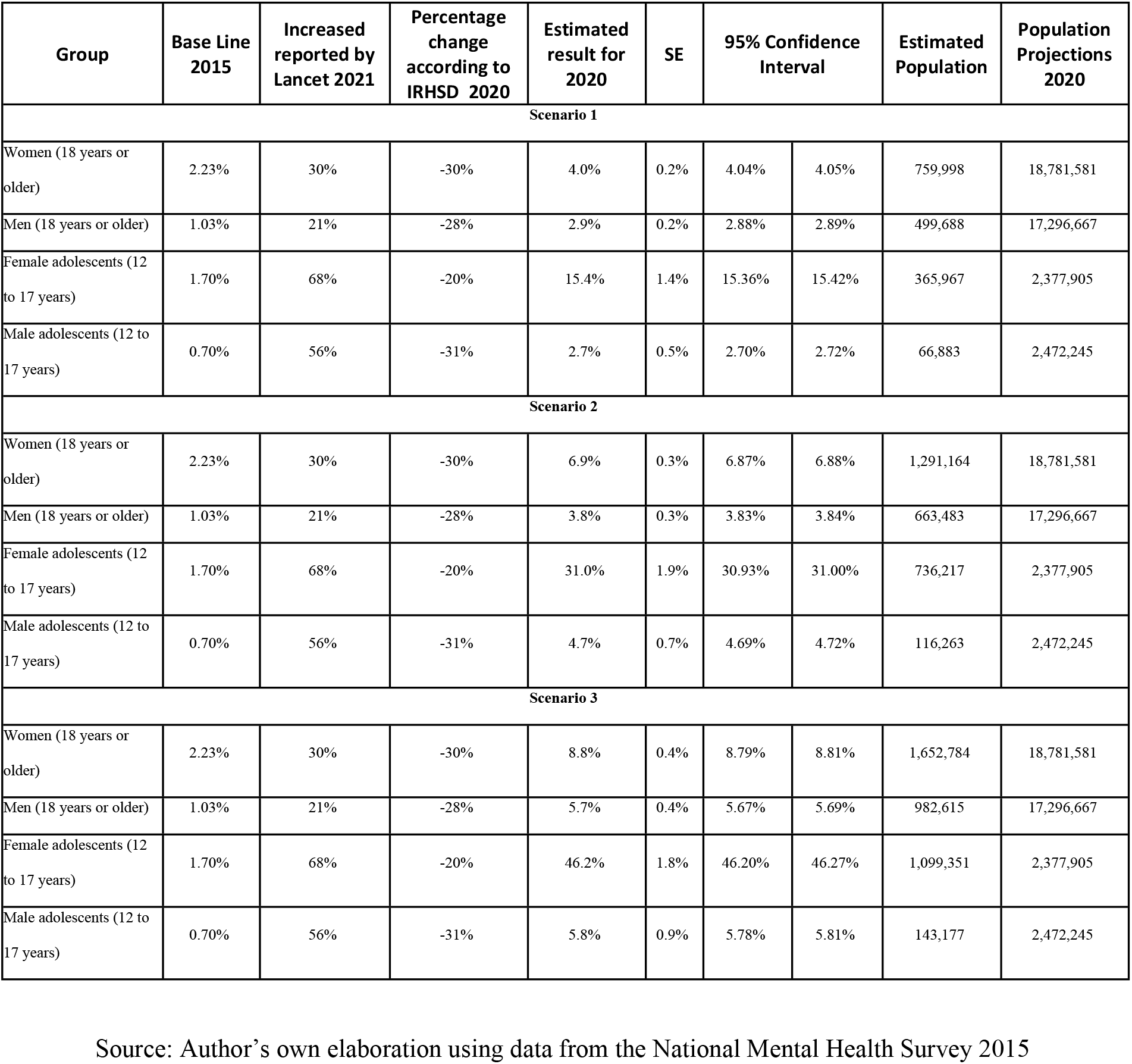
Estimated prevalence of depression for women and men in the three simulated scenarios.

| Group | Base Line 2015 | Increased reported by Lancet 2021 | Percentage change according to IRHSD 2020 | Estimated result for 2020 | SE | 95% Confidence Interval |  | Estimated Population | Population Projections 2020 |
| --- | --- | --- | --- | --- | --- | --- | --- | --- | --- |
| Scenario 1 |  |  |  |  |  |  |  |  |  |
| Women (18 years or older) | 2.23% | 30% | -30% | 4.0% | 0.2% | 4.04% | 4.05% | 759,998 | 18,781,581 |
| Men (18 years or older) | 1.03% | 21% | -28% | 2.9% | 0.2% | 2.88% | 2.89% | 499,688 | 17,296,667 |
| Female adolescents (12 to 17 years) | 1.70% | 68% | -20% | 15.4% | 1.4% | 15.36% | 15.42% | 365,967 | 2,377,905 |
| Male adolescents (12 to 17 years) | 0.70% | 56% | -31% | 2.7% | 0.5% | 2.70% | 2.72% | 66,883 | 2,472,245 |
| Scenario 2 |  |  |  |  |  |  |  |  |  |
| Women (18 years or older) | 2.23% | 30% | -30% | 6.9% | 0.3% | 6.87% | 6.88% | 1,291,164 | 18,781,581 |
| Men (18 years or older) | 1.03% | 21% | -28% | 3.8% | 0.3% | 3.83% | 3.84% | 663,483 | 17,296,667 |
| Female adolescents (12 to 17 years) | 1.70% | 68% | -20% | 31.0% | 1.9% | 30.93% | 31.00% | 736,217 | 2,377,905 |
| Male adolescents (12 to 17 years) | 0.70% | 56% | -31% | 4.7% | 0.7% | 4.69% | 4.72% | 116,263 | 2,472,245 |
| Scenario 3 |  |  |  |  |  |  |  |  |  |
| Women (18 years or older) | 2.23% | 30% | -30% | 8.8% | 0.4% | 8.79% | 8.81% | 1,652,784 | 18,781,581 |
| Men (18 years or older) | 1.03% | 21% | -28% | 5.7% | 0.4% | 5.67% | 5.69% | 982,615 | 17,296,667 |
| Female adolescents (12 to 17 years) | 1.70% | 68% | -20% | 46.2% | 1.8% | 46.20% | 46.27% | 1,099,351 | 2,377,905 |
| Male adolescents (12 to 17 years) | 0.70% | 56% | -31% | 5.8% | 0.9% | 5.78% | 5.81% | 143,177 | 2,472,245 |

Source: Author's own elaboration using data from the National Mental Health Survey 2015

#### Anxiety

In the case of anxiety, it is observed that if we only consider the increase observed in IRHSD (scenario 1), the prevalence of anxiety in 2020 would have been 12.6% for female adolescents, 4.9% for adult women, 4.0% for male adolescents, and 3.4% for adult men.

These prevalences assume that the decrease in the prevalence observed in all groups was the real prevalence for 2020. However, if we include the estimated potential impact of the COVID-19 pandemic on the increase of anxiety, we obtain that the prevalence for female adolescents might reach 23.8%, which corresponds to more than 565,000 female adolescents with anxiety in Colombia (scenario 2). Although the increase for the other groups would not be as large as for female adolescents, in the case of adult women, the prevalence would have increased almost two percentage points between both scenarios, which correspond to almost 300,000 adult women more. Finally, the results of scenario 3 show that the prevalence of anxiety for female adolescents would be 32.4%, followed by male adolescents, with a prevalence of 9.0%. These numbers correspond to more than 220,000 individuals aged 12 to 17 years with anxiety in Colombia.

In the case of adult women, the difference between the second and third scenario is less than one percentage point; however, this represents more than 150,000 women with anxiety. Finally, male adults would have been the group with the lowest increase of the prevalence of anxiety among scenarios. However, under scenario 3, the prevalence for this group might be equal to 6.2%, which correspond to almost 770,000 male adults in the country (Table 3).

### Depression

When we analysed the different scenarios on how the prevalence for depression changed in 2020 and 2021 as a result of the COVID-19 pandemic. We found that when we only use the observed data to estimate the prevalence, the prevalence of depression until 2019 presented an increment in all groups compared to 2015. However, in 2020 the prevalence of depression decreased in all groups, with female adolescents as the group with the highest reduction (- 20%) compared to 2019. Even when considering this reduction, the prevalence of depression for female adolescents had the greatest increase going from 1.7% in 2015 to 15.39% in 2020. which correspond to more than 365.000 female adolescents in the country (scenario 1).

When we analysed scenarios 2, we found that there was an increase in the number of individuals living with a mental health disorder. For example, according to Santomauro (12) it is estimated that the prevalence of depression increased by at least 21% (male adults) and as much as 68% in female adolescents. The results of scenario 2, where we assumed that the increase in the prevalence of depression presented by Santomauro (12) was the real prevalence, we obtained that as expected the increase in the prevalence in depression was greater for female adolescents (31%), followed by adult women (6.9%) and male adolescents (4.7%).

Finally. when we analysed the results of the simulation including the average relative change of the prevalence of depression between 2016 and 2019, assuming that the prevalence of depression would have been like to the one calculated by Santomauro (12) (the additional effect of the COVID-19 pandemic), we found that around 46.2% of female adolescents and almost 9% of adult women might have depression. This represents an increase of 30.8 percentage points compared to scenario 1 and more than 733,000 women (Table 4).

## Discussion

This study aimed to estimate the prevalence of anxiety and depression for female and male adolescents and adults in Colombia before the COVID-19 pandemic and, it also aimed to estimate the potential increase of the prevalence in each group as a result of the COVID-19 pandemic in 2020. To reach this objective we used two data sources; the IRHSD which is the main healthcare administrative record that allows the analysis of the morbidity cases and the National Mental Health Survey for 2015.

The results of this analysis using the IRHSD revealed an important increase in the observed prevalence of these disorders for adults and adolescents and men and women between 2015 and February 2020. However, between March and August 2020, the number of cases decreased, aspect that might be associated with the barriers that individuals faced to have access to different health services as a result of restrictions and lockdown during this period (17). In addition, when we simulated different scenarios using the National Mental Health Survey and estimated the prevalence of both depression and anxiety for adults and adolescents, we found that the prevalence of depression and anxiety increased significantly in the last five years for all groups. This increase has been greater for women than for men, and for adolescents than adults. Depending on the scenario and the assumptions that were made regarding the potential effect of the COVID-19 pandemic on the prevalence of depression and anxiety the results can vary for each of the four groups. For example, in the case of female adolescents the prevalence of depression and anxiety can be as high as 46.2% and 32.4% respectively.

Colombia has followed recommendations by WHO to design national mental health policies and plans (30) nonetheless, the increase in the number of cases for anxiety and depression was only observed after the publication of the National Mental Health Policy in 2018 (31). However, it is not possible to conclude that the increase of cases has been the result of this policy, it can be due to a better registration of patients who consult a physician because of mental health symptoms.

It is important to keep in mind the context of the armed conflict in Colombia and how it has affected the provision of mental health services. Indeed, in 2011 the program of psychological care and integral health for victims of the conflict was created. Its main objective was to mitigate the psychosocial effects of the armed conflict (32). Nevertheless, different studies have revealed the many barriers that victims of the conflict face to access healthcare services. Although, this program exists, little attention has been given to the effect of the conflict on the mental health of non-victims; for example, the National Strategy of Mental Health does not mention the effect of the conflict on mental health, and only refers to other situations and context (33).

The COVID-19 pandemic brought different social, economic, and health challenges. including the provision of healthcare services for non-COVID-19 related diseases. According to the WHO, 75% of the countries reduced the provision of healthcare services or even stopped completely (8) and so was in Colombia (34). In addition, there was an increase in the number and type of barriers that individuals to access healthcare services, starting with the mandatory quarantine that was established between March and August 2020. These barriers might be one of the main reasons why the number of cases diagnosed with depression and anxiety observed during 2020 decreased compared to 2019. However, in April 2020 a mental health hotline was created by the Ministry of Health and Social Protection. During 2020, more than 23,000 interactions of teleconsultation were given to people of all ages and sexes. The principal reasons for consultation were psychological support in stressful situations, anxiety, depression, violence, and suicidal behaviour.

According to our results, women and female adolescents are the groups with the highest number of cases since 2016. However, in 2020, in comparison with men and male adolescents, the differences in the number of cases are greater, reflecting that these groups have a higher need and also look for services more often. This is a similar finding to the one of the global burden of disease and risk factors, which revealed that the burden of depression is 50% higher for females than for males (35). Depression is a common psychiatric disorder, and it is a common mental health problem among adolescents worldwide. In addition, during the COVID-19 pandemic female adolescents were the group with the highest increase in mental health disorders (36). In adolescents, the COVID-19 pandemic was a real challenge as a result of isolation, lack of daily routines, lack of access to health services and schools, and also the lack of capabilities of resilience (37). In addition, periods without school are associated with decreased physical activity, more screen time, irregular sleep patterns (11). The isolation may have an influence on <u>psychiatric disorder</u> onsets during adolescence. For some adolescents, the numerous deaths related to COVID-19-was their first experience with death, and home confinement was associated with an increase in intrafamily violence (38, 39).

Finally, the three scenarios estimated in this paper aimed to explore how the prevalence of depression and anxiety might have increased since 2015 and as a result of the COVID-19 pandemic. Unfortunately, only the National Mental Health Survey allows the estimation of the prevalence of mental health disorders such as depression and anxiety. Therefore, there is not an updated estimator of the prevalence of this illness and more importantly of the characteristics of individuals who are living with depression and anxiety. The results of the three scenarios suggest a potential increase in the prevalence for the four groups (male and female adolescents, and male and female adults). In all groups, we have seen an increase in the number of consultations in the last four years, which means that the prevalence has also increased; Even though there are differences between sex and age groups. For example, female adolescents and female adults are the two groups with the highest increase over the years; this is a common finding in the literature where women of all ages have a higher prevalence of depression and anxiety.

In Colombia there are 2.5 psychiatrists per 100,000 inhabitants that is far from WHO recommendations of 10 per 100,000. In psychology professionals, there are 11 per 100,000 inhabitants. Based on our findings, we strongly recommend strengthening policies, plans, and programs to promote good mental health and early detection of mental health disorders at primary care level with multidisciplinary teams.

### Strength and Limitations

The results of this study contribute to the analysis and understanding of the mental health situation of individuals aged 12 years or older in Colombia. However, it is important to read the results carefully given data limitations. related to the IRHSD. As mentioned before, IRHSD aims to support the payment of health providers; therefore, its main objective is not to collect data on the provision of services. Also, some public providers do not report information to IRHSD given that their payment is made through other systems. Another limitation of the study is the use of the National Mental Health Survey of 2015. Although this is the official source to compute the prevalence of depression, anxiety and other mental health disorders, given that it is seven years old, it might not provide an accurate picture of the current situation of the country.

## Conclusions

Our results suggest that there has been an increase in the number of cases of anxiety and depression since 2016. In addition, our results suggest that the number of cases can increase drastically as a consequence of COVID-19 pandemic. Therefore, the Colombian health system must rethink how mental health services are provided. Also, what strategies are implemented to prevent mental health illness and how to detect potential cases of anxiety and depression, especially in female adolescents. In addition. it is necessary to continue analysing the mental health situation in the country and how it has been affected by the armed conflict of the last six decades.

## Data Availability

The data underlying the results presented in the study are available from the National Mental Health Survey 2015 (http://www.odc.gov.co/Portals/1/publicaciones/pdf/consumo/estudios/nacionales/CO031102015-salud_mental_tomoI.pdf) and the Individual Registry of Health Services Delivery

## APPENDIX 1

### Emergency Cases

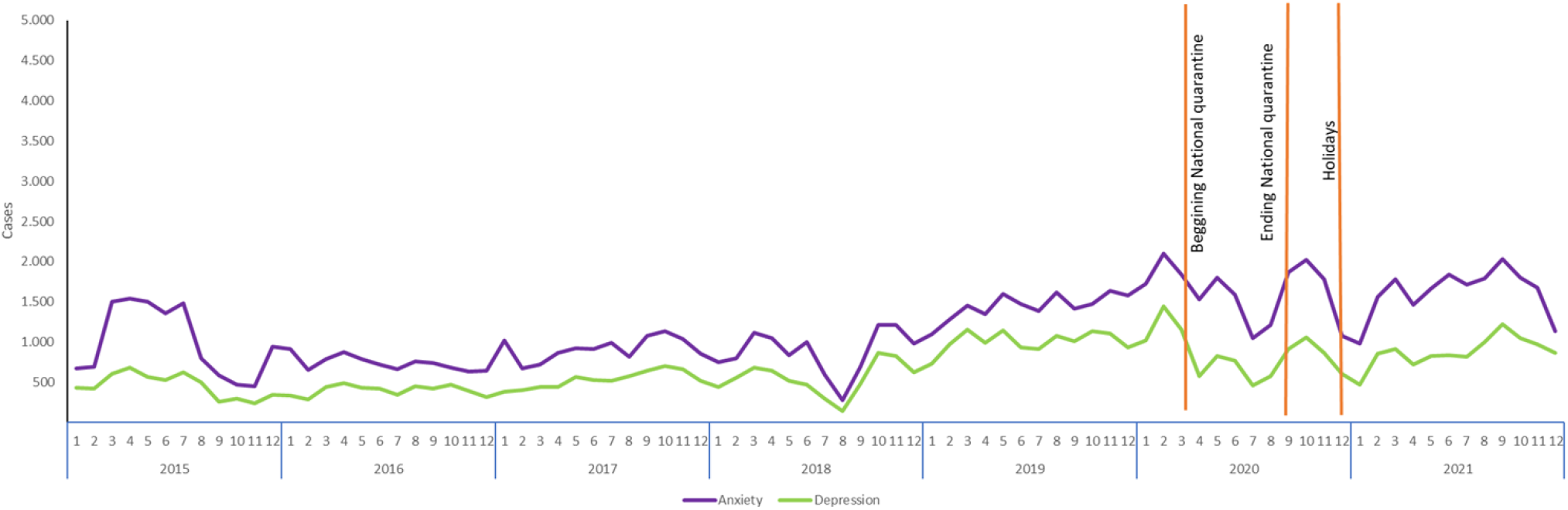

Source: Author’s own elaboration using data from the Individual Registry of Health Services Delivery

### Hospitalization cases

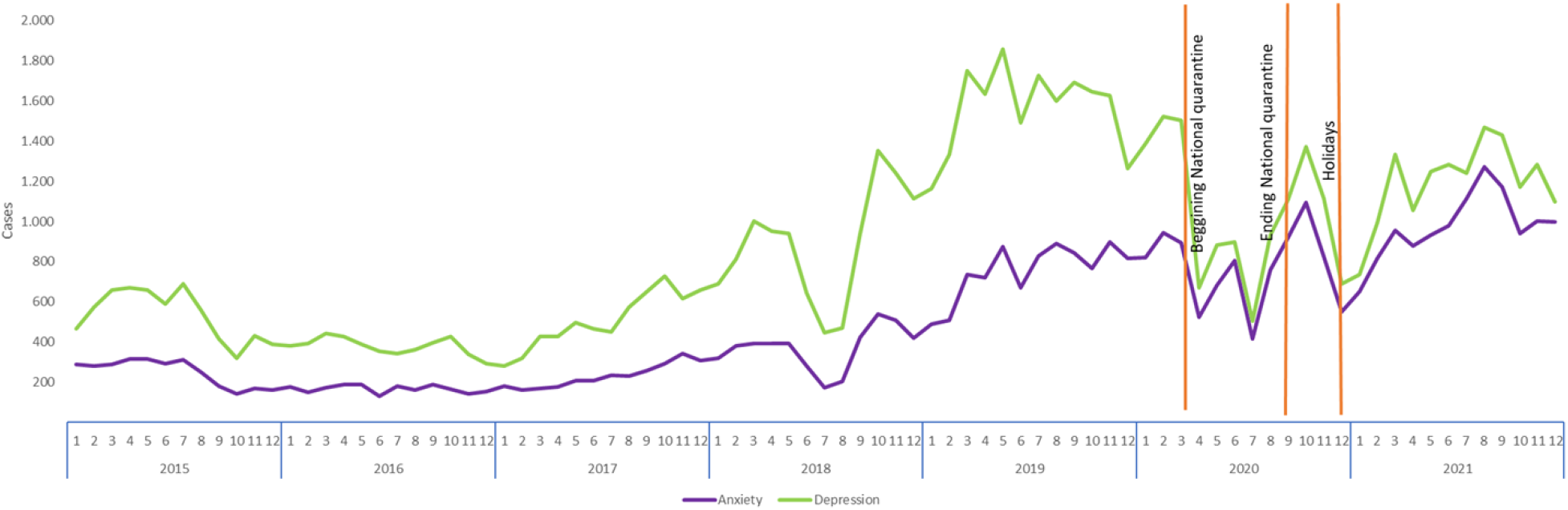

Source: Author’s own elaboration using data from the Individual Registry of Health Services Delivery

#### Comparison cases between adolescents aged 12-17 years by diagnostics

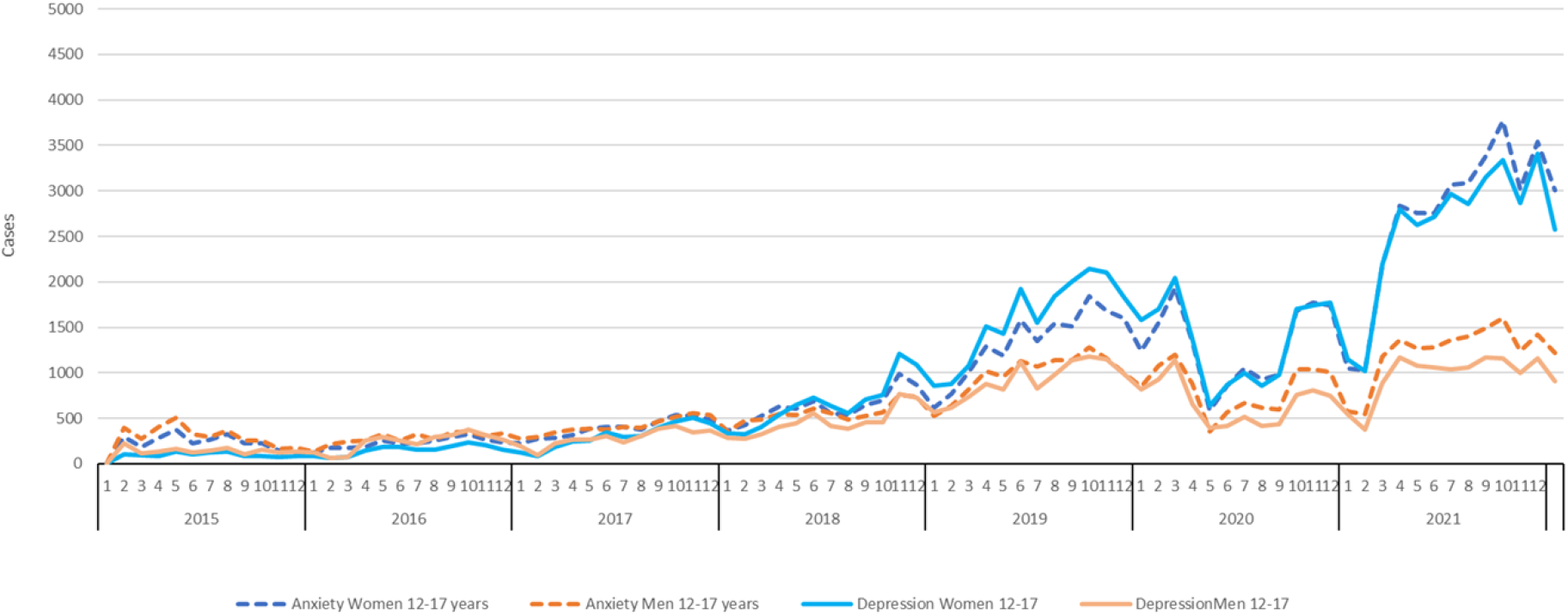

Source: Author’s own elaboration using data from the Individual Registry of Health Services Delivery

#### Comparison cases between adults 18 years or older by diagnostics

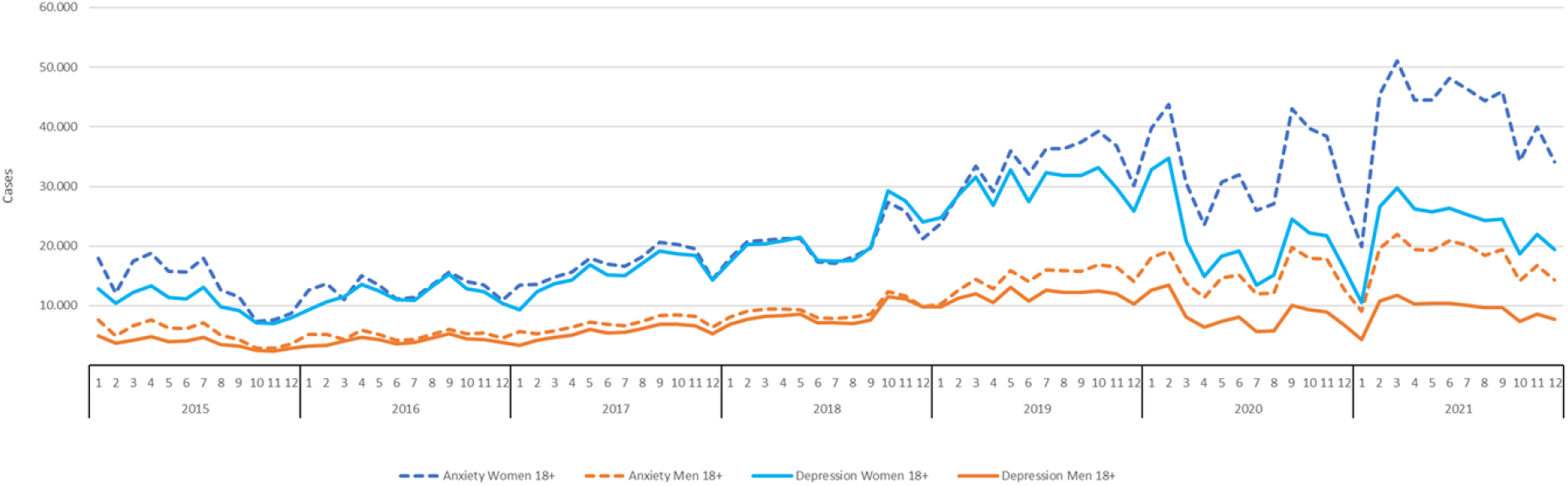

Source: Author’s own elaboration using data from the Individual Registry of Health Services Delivery

## References

1. Else H. The pandemic’s true health cost how much of our lives has COVID stolen. Nature [Internet]. 2022 [cited 2022 Aug 18];605:410–3. Available from: https://www.nature.com/articles/d41586-022-01341-7

2. Bendau A. Petzold MB. Pyrkosch L. Mascarell Maricic L. Betzler F. Rogoll J. et al. Associations between COVID-19 related media consumption and symptoms of anxiety. depression and COVID-19 related fear in the general population in Germany. Eur Arch Psychiatry Clin Neurosci [Internet]. 2021;271(2):283–91. Available from: https://doi.org/10.1007/s00406-020-01171-6

3. Özdin S. Bayrak Özdin Ş. Levels and predictors of anxiety. depression and health anxiety during COVID-19 pandemic in Turkish society: The importance of gender. International Journal of Social Psychiatry. 2020;66(5):504–11.

4. Salari N. Hosseinian-Far A. Jalali R. Vaisi-Raygani A. Rasoulpoor S. Mohammadi M. et al. .????? Global Health. 2020;16(1):1–11.

5. Kola L. Kohrt BA. Hanlon C. Naslund JA. Sikander S. Balaji M. et al. COVID-19 mental health impact and responses in low-income and middle-income countries: reimagining global mental health. Vol. 8. The Lancet Psychiatry. Elsevier Ltd; 2021. p. 535–50.

6. Moreno C. Wykes T. Galderisi S. Nordentoft M. Crossley N. Jones N. et al. How mental health care should change as a consequence of the COVID-19 pandemic. Lancet Psychiatry. 2020 Sep 1;7(9):813–24.

7. Hrynick TA. Ripoll Lorenzo S. Carter SE. COVID-19 response: Mitigating negative impacts on other areas of health. Vol. 6. BMJ Global Health. BMJ Publishing Group; 2021.

8. World Health Organization. COVID-19 disrupting mental health services in most countries. WHO survey [Internet]. 2020 [cited 2022 May 22]. Available from: https://www.who.int/news/item/05-10-2020-covid-19-disrupting-mental-health-services-in-most-countries-who-survey

9. World Health Organization. World Mental Health Day 2020 [Internet]. 2020 [cited 2022 May 22]. Available from: https://www.who.int/es/news/item/27-08-2020-world-mental-health-day-an-opportunity-to-kick-start-a-massive-scale-up-in-investment-in-mental-health

10. Li Y. Scherer N. Felix L. Kuper H. Prevalence of depression. anxiety and posttraumatic stress disorder in health care workers during the COVID-19 pandemic: A systematic review and meta-Analysis. PLoS One. 2021 Mar 1;16(3 March).

11. Wu T. Jia X. Shi H. Niu J. Yin X. Xie J. et al. Prevalence of mental health problems during the COVID-19 pandemic: A systematic review and meta-analysis. Vol. 281. Journal of Affective Disorders. Elsevier B.V.; 2021. p. 91–8.

12. Santomauro DF. Mantilla Herrera AM. Shadid J. Zheng P. Ashbaugh C. Pigott DM. et al. Global prevalence and burden of depressive and anxiety disorders in 204 countries and territories in 2020 due to the COVID-19 pandemic. The Lancet. 2021 Nov 6;398(10312):1700–12.

13. World Health Organization. COVID-19 pandemic triggers 25% increase in prevalence of anxiety and depression worldwide [Internet]. 2022 [cited 2022 May 22]. Available from: https://www.who.int/news/item/02-03-2022-covid-19-pandemic-triggers-25-increase-in-prevalence-of-anxiety-and-depression-worldwide

14. United Nations Development Programme. Human Development Report [Internet]. BERNAN PRESS; 2022 [cited 2022 Sep 18]. Available from: https://hdr.undp.org/system/files/documents/global-report-document/hdr2021-22pdf_1.pdf

15. Panamerican Health Organization. Reporte de situación COVID-19 Colombia No. 252 - 28 de Diciembre 2021 [Internet]. 2021 [cited 2022 May 22]. Available from: https://www.paho.org/es/documentos/reporte-situacion-covid-19-colombia-no-252-28-diciembre-2021

16. Ministerio de Salud y Protección Social. Resolución No. 536 de 2020 [Internet]. 2020. Available from: https://www.minsalud.gov.co/Normatividad_Nuevo/Resoluci%C3%B3n%20No.%20536%20de%202020.pdf

17. Ministerio del Interior. Republica de Colombia. Decreto Número 457 [Internet]. Decreto Número 457 2020 p. 1–14. Available from: https://dapre.presidencia.gov.co/normativa/normativa/DECRETO457DEL22DEMARZODE2020.pdf

18. Ministerio de Salud y de Protección Social. Encuesta Nacional de Salud Mental [Internet]. Bogotá; 2015. Available from: http://www.odc.gov.co/Portals/1/publicaciones/pdf/consumo/estudios/nacionales/CO031102015-salud_mental_tomoI.pdf

19. Cifuentes-Avellaneda Á. Rivera-Montero D. Vera-Gil C. Murad-Rivera R. Sánchez SM. Castaño LM. et al. Informe 3. Ansiedad. depresión y miedo: impulsores de la mala salud mental durante el distanciamiento físico en Colombia [Internet]. 2020 [cited 2022 May 22]. Available from: https://profamilia.org.co/wp-content/uploads/2020/05/Informe-3-Ansiedad-depresion-y-miedo-impulsores-mala-salud-mental-durante-pandemia-Estudio-Solidaridad-Profamilia.pdf

20. Ocampo González ÁA. Castillo García JF. Pabón Sandoval LC. Tovar Cuevas JR. Hidalgo Ibarra SA. Calle Sandoval DA. et al. Depressive symptomatology in adults during the COVID-19 pandemic. J Investig Med. 2022 Feb 1;70(2):436–45.

21. Monterrosa-Castro Á. Monterrosa-Blanco A. González-Sequeda A. Perceived Loneliness and Severe Sleep Disorders in Adult Women during the Covid-19 Quarantine: A Cross-Sectional Study in Colombia. J Prim Care Community Health. 2021;12.

22. Sanabria-Mazo JP. Useche-Aldana B. Ochoa PP. Rojas-Gualdrón DF. Sanz A. Estudio PSY-COVID Impacto de la pandemia de COVID-19 en la salud mental en Colombia [Internet]. 2021 [cited 2022 May 22]. Available from: https://www.colpsic.org.co/wp-content/uploads/2021/09/Libro-Impacto-de-la-pandemia-de-COVID-19-en-la-salud-mental-en-Colombia.pdf

23. Departamento Nacional de Planeación. ¿Cómo se relaciona la pandemia del COVID-19 con la salud mental de los colombianos? [Internet]. 2021 [cited 2022 May 10]. Available from: https://colaboracion.dnp.gov.co/CDT/Sinergia/Documentos/Notas_politica_publica_SALUD%20MENTAL_22_04_21_V7.pdf

24. Gonzalez-Uribe C. Olmos-Pinzon A. Leon-Giraldo S. Bernal O. Moreno-Serra R. Health perceptions among victims in postaccord Colombia: Focus groups in a province affected by the armed conflict. PLoS One. 2022 Mar 1;17(3 March).

25. Rojas Y. Torres C. Figueredo M. Hernández F. Castañeda C. Lasalvia P. et al. Estimación de la prevalencia de EPOC en Colombia a partir del Registro Individual de Prestaciones de Servicios de Salud. Revista Colombiana de Neumología [Internet]. 2019;31(1):5–15. Available from: http://dx.doi.org/10.30789/rcneumologia.v31.n1

26. Ministerio de Salud y de Protección Social. Preguntas frecuentes RIPS ¿Qué son los RIPS? [Internet]. 2015. p. 1–17. Available from: https://www.minsalud.gov.co/sites/rid/Lists/BibliotecaDigital/RIDE/DE/OT/FAQ-RIPS.pdf

27. Departamento Administrativo Nacional de Estadística. Proyecciones de Población [Internet]. [cited 2022 Jun 5]. Available from: https://www.dane.gov.co/index.php/estadisticas-por-tema/demografia-y-poblacion/proyecciones-de-poblacion

28. García-Alonso CR. Almeda N. Salinas-Pérez JA. Gutiérrez-Colosía MR. Uriarte-Uriarte JJ. Salvador-Carulla L. A decision support system for assessing management interventions in a mental health ecosystem: The case of Bizkaia (Basque Country. Spain). PLoS One. 2019 Feb 1;14(2).

29. Torres-Jiménez M. García-Alonso CR. Salvador-Carulla L. Fernández-Rodríguez V. Evaluation of system efficiency using the Monte Carlo DEA: The case of small health areas. Eur J Oper Res. 2015 Apr 16;242(2):525–35.

30. Henao S. Quintero S. Echeverri J. Hernández J. Rivera E. López S. Políticas públicas vigentes de salud mental en Suramérica: un estado del arte. Revista Facultad Nacional de Salud Pública [Internet]. 2016 May 2;34(2). Available from: https://revistas.udea.edu.co/index.php/fnsp/article/view/23468

31. Ministerio de Salud y Protección Social. Política Nacional de Salud Mental [Internet]. 2018 [cited 2021 Jul 4]. Available from: https://www.minsalud.gov.co/sites/rid/Lists/BibliotecaDigital/RIDE/VS/PP/politica-nacional-salud-mental.pdf

32. Ministerio de Justicia. Decreto 4800 de 2011. Colombia; 2011.

33. Departamento Nacional de Planeación. Estrategia para la promoción de la salud mental en Colombia. CONPES 3992. cOLOMBIA; 2020.

34. Banco Mundial. Evaluación de las capacidades de preparación y respuesta ante futuras pandemias y emergencias en salud pública [Internet]. 2022. Available from: http://www.worldbank.org

35. Lopez AD. Mathers CD. Global Burden of Disease and Risk Factors. Washington: Oxford University Press and the World Bank; 2006.

36. Zhou SJ. Zhang LG. Wang LL. Guo ZC. Wang JQ. Chen JC. et al. Prevalence and socio-demographic correlates of psychological health problems in Chinese adolescents during the outbreak of COVID-19. Eur Child Adolesc Psychiatry. 2020;29(6).

37. Jones EAK. Mitra AK. Bhuiyan AR. Impact of covid-19 on mental health in adolescents: A systematic review. Int J Environ Res Public Health. 2021 Mar 1;18(5):1–9.

38. Guessoum SB. Lachal J. Radjack R. Carretier E. Minassian S. Benoit L. et al. Adolescent psychiatric disorders during the COVID-19 pandemic and lockdown. Psychiatry Res. 2020 Sep 1;291.

39. de Figueiredo CS. Sandre PC. Portugal LCL. Mázala-de-Oliveira T. da Silva Chagas L. Raony Í. et al. COVID-19 pandemic impact on children and adolescents’ mental health: Biological. environmental. and social factors. Prog Neuropsychopharmacol Biol Psychiatry. 2021 Mar 2;106.

